# Intentions and willingness to engage in risky driving behaviour among high school adolescents: evaluating the bstreetsmart road safety program

**DOI:** 10.1101/2022.03.05.22271967

**Authors:** LN Sharwood, A Martiniuk, P Sarrami, J Seggie, S Wilson, J Hsu, B Burns, D.B. Logan

## Abstract

**Objective:** investigate the impact of a road-safety program on adolescent willingness to engage in risky behaviour as probationary drivers, adjusted for covariates of interest.

**Method:** *b*streetsmart is a road-safety program delivered to around 25,000 adolescent students annually in New South Wales. Using a smart phone-based app, student and teacher participation incentives, students were surveyed before and after program attendance. Mixed methods linear regression analysed pre-post modified Behaviour of Young Novice Driver (BYNDS_M) scores.

**Results:** 2360 and 1260 students completed pre- and post-event surveys respectively. Post-event BYNDS_M scores were around 3 points lower than pre-event scores (−2.99, 95%CI - 3.418 to -2.466), indicating reduced intention to engage in risky driving behaviours. Covariates associated with higher stated intentions of risky driving were exposure to risky driving as a passenger (1.21, 95% CI 0.622-2.011), identifying as non-binary gender (20.8, 95% CI 8.795 to 40.852), adjusting for other predictors.

**Conclusions:** Trauma-informed, reality-based injury prevention programs are effective in changing short term stated intentions to engage in risky driving, among a pre-independent driving student population. The adolescent novice driver age group is historically challenging to engage, and injury prevention action must be multi-pronged to address the many factors influencing their behaviour.

**What is already known on this topic:** Road traffic injuries are the leading cause of death for adolescents in most developed countries globally. Injury prevention action must be multi-pronged to address the many factors influencing their behaviour

**What this study adds:** The bstreetsmart injury prevention intervention which incorporates trauma informed, CBT influence and reality-based road safety information to around 25,000 students annually, showed significant short-term impact on the stated willingness of the study population to engage in risky driving behaviour when obtaining their probationary licence. Adolescents are strongly influenced by examples of risky road behaviours among their closest adult drivers.

**How this study might affect research, practice, or policy:** Interventions such as bstreetsmart hold a positive place in the multi-pronged approach needed to address the difficult issue of novice drivers.

## Introduction

Serious injuries and road deaths among young novice drivers remain unacceptably high and are a serious public health issue. These road users have some of the highest crash rates in Australia; drivers aged 16-19 years are 6-8 times more likely to crash than those aged 55-59 years^1^ and a young novice driver on probationary first year license in Australia is four times more likely to be involved in a fatal crash than a 26 year-old driver^2^. Young novice driver crash and fatality rates are highest in the first year, peaking immediately post-licensure^3^. This pattern is similar worldwide^4^, contributing to the fact that road traffic injuries are the leading cause of death for adolescents in most developed countries globally^5^.

Young drivers either underestimate the complexity of the driving task, overestimate their skill, or both; the consequence imposing a smaller safety margin than they perceive exists^6 7^. Factors influencing novice driver risk include age, gender, maturity^8^, knowledge and attitudes, motivation, and sensation-seeking^4^. These factors impact involuntary and voluntary risky driver behaviour and are highly challenging when developing effective countermeasures. Some research has identified a relationship between the perceived risky driving behaviour of parents and friends and the probationary license holder’s willingness to engage in risky behaviour themselves^6 9^. This highlights a need to broaden education beyond the pre-licensed driver, aiming to influence driving behaviour related influences they may have at home or among peers. However, further information regarding these relationships is needed.

Gender differences among young drivers have been predominantly investigated using binary (i.e., male/female) attributed populations, showing males to be more willing to take driving risks^7^, have more traffic violations^7 10^ and higher fatality rates than females^1^. Sexual minority adolescents (e.g., identifying as non-binary gender), have self-identified as engaging in higher rates of risky health behaviours such as drug/alcohol and intentional self-harm compared with heterosexual adolescents^11 12^, yet no peer-reviewed evidence has been found on the relationship between sexual minority adolescents and risky driving behaviours.

Behaviour change techniques (BCT) have been successfully employed to change a variety of health behaviours, such as weight loss. BCTs are defined as observable, replicable component of an intervention that is designed to change behaviour^13^. There have been many initiatives in recent years addressing novice driver behaviour such as graduated licensing schemes, education and training programs for adolescents and their parents, legal enforcements^14^, and psychosocial education interventions aiming to impact upon sensation seeking and risk-taking intentions. However, there is little consensus on which approaches, if any, are effective in achieving lasting change in road-user behaviour and relatively few that are theory-led or evidence-based^15^. Michie et al^13^ described the concept of willingness to perform a target behaviour which is impacted upon by proposed key concepts for successful BCT such as ‘shaping knowledge’ (assisting BCT recipients to better understand their behaviour), ‘social comparison’ (generally comparing to positive examples of a desired behaviour) and ‘comparison of outcomes’ (allowing exploration of the outcomes of exhibiting or not exhibiting the behaviour). Some of these BCT concepts are incorporated into the ***b***streetsmart initiative.

‘***b***streetsmart’ is an injury prevention program encouraging safer road-user behaviours among New South Wales (NSW) adolescents, established in 2005 by Westmead Hospital Trauma Department, NSW, Australia (https://bstreetsmart.org/). From the ∼450 students witnessing the inaugural event, ***b***streetsmart has grown to around 25,000 pre-driving adolescents from NSW schools attending ***b***streetsmart annually, over a 3-day period. Students witness crash re-enactments, potential risk-taking consequences, engage in interactive displays, hear from young drivers permanently injured in road crashes, and learn injury prevention strategies. ***b***streetsmart’s primary objective is to reduce the fatality and injury rates of young people by promoting safe behaviour; offering students and teachers access to current information injury prevention strategies. Its development has involved significant consultation with NSW Ambulance Service, Fire and Rescue NSW, NSW Police Force and NSW Police Crash Investigation Unit, Transport for NSW and NSW High School Year Advisors. ***b***streetsmart has undergone several process evaluations (none published), all suffering from small response fractions (< 5%) from students and teachers. However, its capacity to influence the future behaviour of the adolescents transitioning into driving remains unknown.

The Behaviour of Young Novice Drivers Scale (BYNDS) was developed in 2010^16^ and has been validated since across various populations^17-19^. The BYNDS intended investigation of the underlying dimensions to young novice driver risky behaviour (i.e., probationary drivers), however the authors deemed it equally appropriate to measure stated intentions regarding risk among pre-independent driving youth. Factors measured within the full BYNDS scale comprised those deemed *transient* that would occur during a driving episode (e.g. speeding), those taken prior to a driving episode classified as *fixed* for within journey risk (e.g. drinking alcohol before driving), risky driving exposure (e.g. driving drunk friends) and those measuring driver emotions or *mood* prior to or during a journey. Substantial research describes adolescent development and the impulsivity common among this age group, typical of a young person in the process of creating their identity, values, and opinions^20^. During this phase, adolescents are deemed more sensitive to social influences and role modelling, and normative aspects of their environment have been shown to assert strong influence over a novice driver’s intentions^4 20^. This is described as the young person’s ‘social comparison’, where they may be more likely to do what they think others do. A version of the BYNDS for passengers (BYNDS_P) was developed by Dr Bridie Scott-Parker while conducting this study, to assess the risky driving behaviour of parents and friends with whom a pre-driving adolescent was a passenger (unpublished). The BYNDS_P was included in the pre-event survey to additionally understand the contribution of normative aspects of an adolescent’s road experience and risky driving exposure, to their own stated intentions of willingness to engage in risky driving behaviour.

The aim of this study was to explore self-reported willingness to engage in risky driving behaviour using a validated measure, and the influence of a range of covariates including gender, socio-economic status, and exposure to risky driving behaviour (BYNDS_P). We aimed to explore whether the ***b***streetsmart event could influence adolescents’ stated intentions to engage in risky driving.

## Method

### Participants

A pre-post longitudinal survey was designed with three time points for data collection: the first was prior to attendance at the September 2019 ***b***streetsmart event, the second a follow-up response within the week after ***b***streetsmart attendance, and finally a second follow-up at 3 months. An electronic study information pack was embedded within a mini-site (web-based ‘app’) accessible via smart-phone devices, tablet or personal computer. Participant consent was sought within the online survey, with data privacy assurance and participant information provided prior to their completion of survey responses. Ethics approval permitted pre-event student contact via their registered schools, comprising one of the 202 NSW public, private and independent secondary schools that signed on for the event. Attending students were also given the option of accessing the app and providing informed consent to sign up and complete the survey prior to the event commencement. Two researchers attended the event site on each of the three days, to provide students with the relevant information regarding participation and incentives before they entered the arena.

Ethical approval was obtained to develop a socio-economic, health and wellness and behavioural profile of participating students (University of Sydney Human Research Ethics Committee, 2019/510). The approved and internationally validated screening tools included the Kessler Psychological Distress Scale^21 22^ as a mental health screen of the study population, and the Brief Sensation Seeking Scale^23 24^ (first published in 2002) to identify behaviours including attraction to high-risk activities, self-reported health conditions, hours of sleep and substance use including alcohol and drugs. Several elements of these tools needed to be removed at the request of the NSW Education Research department who preferred parental consent. Due to the timeframes of this project, parental consent was not able to be sought.

Participants were incentivised with a chance to win one of three new Apple iPhone XS smart phones provided they completed both the pre and post event surveys, with 50 movie vouchers also available upon completion of the 3–6-month follow-up survey. Teachers were incentivised for their time with the chance to win science incursions (Fizzics Education™) in informing their students of the study; eligible to enter a draw to win one of three science incursions if at least 50% of their attending students completed the surveys. For students without access to their own smart device, teachers facilitated access during class time on school devices.

The pre-event survey requested basic demographic information, including age (years), gender (male, female, non-binary), living situation (i.e., living at one or between two houses), residential postcodes, cultural background, and learner permit attainment and driving and incident exposure. Primary residential postcodes enabled generation of Index of Relative Socio-economic Advantage and Disadvantage, a summary measure within the 2016 Socio-Economic Indexes for Areas (SEIFA)^25^. SEIFA quintiles were used identifying areas of residence from the most socioeconomic advantaged (quintile 5) to the least advantaged (quintile 1). Identification of cultural and ethnic groups adhered to the Australian Standard Classification of Cultural and Ethnic Groups 2019^26^.

The outcome variable used was the modified Behaviour of Young Novice Drivers Scale (BYNDS_M)^16^ which determined adolescents’ stated intentions to engage various behaviours once on probationary licences. The BYNDS_M employed 16 of the original 44 items, including five items assessing *transient* violations, six assessing *fixed’* violations and five assessing *mood*. Scott-Parker^16 18^ approved reduced response levels to enable timely survey completion; BYNDS_M Likert scale responses restricted to - ‘Never’, ‘Sometimes’ or ‘Nearly all the time’, instead of the original five responses^16^. Lower scores (minimum of 16) indicated less or no intention to engage in risky driving, higher scores the opposite (maximum of 48).

The ‘passenger BYNDS’ (BYNDS_P) was used to describe exposures to risky driving behaviours while a passenger with ‘the driver on most journeys over the past month’. Nine statements also requested the same three Likert scale responses from which a variable ‘risk exposed’ was generated. Lower scores interpreted as no or low risk of exposure to risky driving (minimum=9), and higher scores the inverse, interpreted as high or maximum risk of exposure to risky driving (maximum=21). Follow up surveys repeated the BYNDS_M to gauge any change in perceptions of stated intentions to engage in risky behaviour after attending ***b***streetsmart.

### Statistical analyses

Survey data was imported from the online database to conduct analysis using STATAv16 (STATACorp/v16.1/IC) and RStudio. Summary statistic reporting followed internationally standardised formats; parametric data described using mean and standard deviations, p-values of association significant at <0.05. Group means were compared using t-tests. Linear regression was used to analyse pre-event survey data to identify covariates significantly associated with BYNDS_M score outcomes. Potential predictors included in the base model were age, gender, SEIFA quintile score, BYNDS_P, learner permit and cultural background. Model selection was conducted using the ‘step’ function in R-base package, which reduces the model by comparing all possible models sequentially, evaluating using Akaike’s information criterion (AIC). A linear mixed-model regression analysis was then conducted to compare results from the pre and post surveys, using a within subjects design with a random effect for participant, against the outcome variable of the BYNDS_M score.

Ethics approval was obtained from both University of Sydney HREC (2019/510) and State Education Research Applications Process (SERAP)(DOC19/720207/SERAP2019347). This study was reported using the TIDieR checklist.

## Results

A total of 2360 secondary school students consented to participate in the pre-event survey, 61% were female and 4.0% Indigenous. A total of 25,047 students from 208 secondary schools had registered to attend the event. Table 1 describes characteristics of the study population.

**Table 1:** Characteristics of the baseline population (n=2360)

| Characteristic | N (%) |
| --- | --- |
| Age – mean (SD) | 15.7 (0.68) |
| Gender |  |
| ▪ Female | 1432 (60.7) |
| ▪ Male | 868 (36.8) |
| ▪ Non-binary | 60 (2.5) |
| Indigenous (=yes) | 95 (4.0) |
| Cultural background |  |
| ▪ North African/Middle Eastern | 241 (10.1) |
| ▪ Northeast Asian | 85 (3.6) |
| ▪ Northwest European | 328 (13.9) |
| ▪ Oceanian | 930 (39.4) |
| ▪ Americas | 53 (2.2) |
| ▪ Southeast Asian | 310 (13.1) |
| ▪ South/Central Asian | 95 (4.0) |
| ▪ South/Eastern European | 277 (11.7) |
| ▪ Sub-Saharan African | 36 (1.5) |
| ▪ Missing | 8 (0.3) |
| Live between two houses (=yes) | 337 (14.3) |
| SEIFA score (primary) |  |
| ▪ 5 (most advantaged) | 957 (40.5) |
| ▪ 4 | 332 (14.1) |
| ▪ 3 | 405 (17.2) |
| ▪ 2 | 239 (10.1) |
| ▪ 1 (least advantaged) | 427 (18.1) |
| Have obtained Learner permit |  |
| ▪ Yes | 932 (39.5) |
| ▪ No | 1404 (59.5) |
| ▪ Had one but now suspended | 16 (0.6) |
| Hours driven on Learner permit median (IQR) | 20 (5, 50) |
| Have crashed on Learner permit | 30 (3.2) |
| ▪ Yes |  |

Of the original study cohort of 2360 students, the outcome variable (the modified BYNDS score) was completed by 2342 students (99.2%). Median imputation completed the missing scores. The 3-6 months post survey was completed by 204 students (8.6%) and was considered too small a sample to be used. The post-event survey was completed by 1260 (53.4%) of the 2360 cohort, for whom survey data were linked (Table 2). Comparisons of summary scores of the outcome variable in the pre- and post-event surveys in Table 2 were achieved using t-tests to compare raw scores in gender sub-groups. In the post-event survey (around one week after the ***b***streetsmart event), females, males and non-binary students showed lower overall scores than prior to the program, indicating lower stated willingness to engage in risky behaviour ‘when I get my P’s’ (probationary license in Australia).

**Table 2:** Summary scores pre- and post-event for the modified Behaviour of Young Novice Driver Scale.

| **Modified BYNDS Score** | **Pre-event**<br>**(N= 2360)** | **Post-event**<br>**(N= 1260)** | **P-value** |
| --- | --- | --- | --- |
| Females [N (%)]<br>- mean (SD) score | 1432 (60.7)<br>18.3 (3.6) | 750 (59.5)<br>16.3 (3.5) | <0.001 |
| Males [N (%)]<br>- mean (SD) score | 868 (36.8)<br>18.8 (4.5) | 482 (38.2)<br>17.4 (4.0) | <0.001 |
| Non-binary [N (%)]<br>- mean (SD) score | 60 (2.5)<br>22.6 (8.7) | 28 (2.2)<br>16.4 (2.2) | <0.001 |
| Total [N (%)]<br>- mean (SD) score | 2360 (100)<br>18.6 (4.2) | 1260 (100)<br>16.7 (3.7) | <0.001 |

The linear regression using the pre-event data and predictor removal using the AIC, removed the cultural background, SEIFA score and learner permit variables. The linear mixed model included: the ‘risk exposed’ variable, age, sex, and interactions between age and sex. Results are shown in Table 3, identifying that taking into account pre-event risk exposure in parental driving, age and sex, the post-event scores were on average three units lower than the pre-event scores (−2.99, 95% CI -3.418 to -2.466). Exposure to risky driving as a pre-driving teen increased stated intentions to engage in risky driving, with higher modified BYNDS score (1.21, 95% CI 0.622 to 2.011), and compared to females (reference), students identifying as non-binary reported significantly higher BYNDS_M scores (20.8, 95% CI 8.795 to 40.852), after adjusting for other predictors.

**Table 3:** Linear mixed regression analysis – final model.

| Fixed effects | Estimate | p-value | 95% CI |
| --- | --- | --- | --- |
| Intercept | 21.88 | <0.001 | 18.049 to 25.730 |
| Time – post-event | -2.99 | <0.001 | -3.418 to -2.466 |
| Risk exposed | 1.21 | <0.001 | 0.622 to 2.011 |
| Age | -0.36 | 0.003 | -0.612 to -0.122 |
| Sex – female | (ref) |  |  |
| Sex - Male | -0.99 | 0.81 | -0.009 to 0.246 |
| Sex – non-binary | 24.8 | 0.002 | 8.795 to 40.852 |
| Interactions |  |  |  |
| Age:MaleSex | 0.072 | 0.71 | -0.317 to 0.461 |
| Age:Sex-non-binary | -1.57 | 0.002 | -2.589 to -0.564 |

Table 4 compares student responses regarding various risky driving behaviours that they quantified in the adult who ‘most commonly drives them around’ (BYNDS_P) with their own stated intentions ‘when I have my probationary license’ (BYNDS_M). Notable differences included intending to “Sometimes drive without a valid licence” (9% versus∼ 2%), “Sometimes drive without a seatbelt” (5.8% versus 2.6%), “Sometimes drive drugged” (2.9% versus 1.1%) and “Sometimes carry too many passengers” (17.7% versus 5.6%) as probationary license holders.

**Table 4:** **Comparison of parent/carer’s driving behaviour and teen’s intentions in pre-survey students**

| Behaviour | The adult who drives me most of the time: N (%) | When I have my P’s I will: N (%) | Chi-squ p-value: |
| --- | --- | --- | --- |
| ▪ Nearly always drive over speed limit<br>▪ Sometimes drive over speed limit<br>▪ Never drive over speed limit<br>▪ Missing | 107 (4.6)<br>789 (33.7)<br>1446 (61.7)<br>0 (0) | 112 (4.8)<br>848 (36.2)<br>1382 (59.0)<br>18 (0.7) | 649.7<br><0.001 |
| ▪ Nearly always drive without a seatbelt<br>▪ Never drive without a seatbelt<br>▪ Sometimes drive without a seatbelt<br>▪ Missing | 135 (5.7)<br>2159 (91.5)<br>61 (2.6)<br>5 (0.2) | 52 (2.2)<br>2141 (90.7)<br>137 (5.8)<br>30 (1.3) | 462.4<br><0.001 |
| ▪ Nearly always drive drugged | 23 (0.9) | 29 (1.2) | 1.5e+.03<br><0.001 |
| ▪ Sometimes drive drugged | 27 (1.1) | 69 (2.9) |  |
| ▪ Never drive drugged | 2306 (97.7) | 2227 (94.3) |  |
| ▪ Missing | 4 (0.7) | 35 (1.4) |  |
| ▪ Nearly always carry too many passengers | 30 (1.2) | 50 (2.1) | 885.7<br><0.001 |
| ▪ Sometimes carry too many passengers | 132 (5.6) | 419 (17.7) |  |
| ▪ Never carry too many passengers | 2194 (92.9) | 1853 (78.5) |  |
| ▪ Missing | 4 (0.2) | 38 (1.6) |  |
| ▪ Nearly always drive without a valid licence | 31 (1.3) | 69 (2.9) | 697.4<br><0.001 |
| ▪ Sometimes drive without a valid licence | 45 (1.9) | 213 (9.0) |  |
| ▪ Never drive without a valid licence | 2279 (96.5) | 2037 (86.3) |  |
| ▪ Missing | 5 (0.2) | 41 (1.7) |  |
| ▪ Nearly always talk with mobile phone in hand while driving | 67 (2.8) | 51 (2.2) | 998.3<br><0.001 |
| ▪ Sometimes talk with mobile phone in hand while driving | 507 (21.4) | 392 (16.6) |  |
| ▪ Never talk with mobile phone in hand while driving | 1773 (75.1) | 1854 (78.5) |  |
| ▪ Missing | 13 (0.5) | 63 (2.7) |  |
| ▪ Nearly always speed when in a bad mood | 61 (2.6) | 60 (2.5) | 1.0e+03<br><0.001 |
| ▪ Sometimes speed when in a bad mood | 599 (25.4) | 620 (26.3) |  |
| ▪ Never speed when in a bad mood | 1692 (71.7) | 1634 (69.2) |  |
| ▪ Missing | 8 (0.3) | 46 (1.9) |  |

## Discussion

This study has demonstrated a significant reduction in the stated intentions of young pre-driving drivers to engage in risky driving behaviour, within the week following attendance at the bstreetsmart event demonstrating risks and consequences of unsafe driving practices. Willingness was measured by the BYND_M, a modification of a validated tool among novice drivers^16^, an important aspect when implementing BCT^13^. The impact appeared most effective among females with younger age, which supports existing literature of young males having higher rates of risky or delinquent behaviour compared with females^27^. The impact appeared less effective among adolescents identifying as non-binary, and those exposed to risky driving behaviours while a passenger.

Behavioural willingness to engage in risk has been previously explored in an Australian driving population aged 17-25 years^28^, willingness measured as a response to risk-related scenarios using the Prototype Willingness Model^29^, and comparing probationary drivers (years 1 and 2) with open driver license types. The authors identified younger drivers with higher behavioural willingness was associated with lower perceived risk and higher reported engagement in risky driving behaviour^28^. They did not, however explore genders other than male and female, nor the influence of other driver modelled behaviour. Gender identity minorities among adolescents are evidenced to have high risk-taking behaviours compared to cis-gender adolescents, for example having higher rates of intentional self-harm including suicide attempt^30^. There are limited data regarding motor vehicle crashes by sexual orientation, and administrative data collections that document options other than male, female or undetermined sex are rare, and sexual identity does not form part of administrative data collections in general. Reisner et al ^31^ explored the impact of sexual minority status on self-reported seat belt use in American adolescents, finding adolescents of sexual minority orientation to have significantly higher rates of safety belt non-use. Their suggestions as to why this might be focussed on the idea that non-conformity to gender norms as both a pathway to sexual minority orientation and personality characteristics. They proposed that this non-conformity could also include rejection of other ‘conformity’ related behaviours such as seatbelt wearing ^31^. Sexual minorities have similarly been shown to report more risky behaviours than their cisgender counterparts for unhealthy behaviours linked with cancers such as alcohol, tobacco and other drug use, sexual activity, and dietary habits. ^27^

The American College of Surgeons Committee on Trauma updated published recommendations for the optimal care of the injured patient ^32^, recommending that “…Trauma centres must implement at least two programs that address one of the major causes of injury in the community”, encouraging trauma services to incorporate injury research and proactive preventive programs into their remit as part of a community. Many programs that have been implemented have focussed on fear appeal approaches, however despite the modest effects of attitudinal change seen in the short term, they have received substantial criticism of the risks of psychological harm with this approach^33 34^. Rather than using ‘shock tactics’ or basing prevention strategies on fear appeals, Purtle et al ^35^recommend trauma-informed programs which acknowledge previous exposures to trauma and offer services that recognise the emotional, social and cognitive impacts of these experiences.

Since its inception in 2005 and due to its popularity, ***b***streetsmart has seen 207,955 students attending to end 2019. The program has more recently been live-streamed, making exposure counts less certain. The trauma-informed approach includes integration of a trauma-informed approach with the crash re-enactment, speakers with brain and spinal injuries and a family member who has lost a teen driver in a road crash. The potential emotional impact on students and teachers is managed by extensive pre-event communications with schools and provision of preparatory material. During the event multiple trained counsellors are available for assistance, debriefing and support if needed.

This study had several strengths including its sample size and reasonable response fraction for this demographic (53.4%). The population was likely to be representative of the NSW student population, given over 200 schools from greater Metropolitan Sydney area participated. Although around 40% of the study population resided in the socio-economically advantaged area, the less advantaged populations were also evenly represented, with almost one fifth of students(18.1%) living in the least socio-economically advantaged areas, according to the SEIFA scores^25^. A limitation of comparing the BYNDS_P with BYNDS_M (Table 4) responses concerning passenger carrying, is the potential influence of known passenger restrictions for probationary drivers only. The main study limitation was the restriction to the amount of data permitted to be collected by the second stage of ethical review, limiting the exploration of what may have been important covariates of the outcome of interest. SERAP’s concerns were that parental consent was not obtained. Recent systematic review exploring adolescent consent ^36^ concluded that most adolescents were able to comprehend the nature and purpose of proposed research, correctly interpret benefits and risks, and thus provide their own consent. Comprehension increased with age and school year level; adolescents aged 15–16 years demonstrating a similar understanding of research to adults.

## Conclusion

This study demonstrates a trauma-informed, reality-based injury prevention program as effective in changing short-term stated intentions to engage in risky driving, among pre-independent driving students. Non-binary identifying students showed higher intentions to engage in risky driving and prevention programs should be responsive to these tendencies among sexual minority adolescents. These findings should encourage ongoing research in this field and the continued efforts of prevention programs targeted toward pre-driving adolescents.

## Data Availability

All data produced in the present study are available upon reasonable request to the authors

## Acknowledgements

We gratefully acknowledge funding from the Institute for Trauma and Injury Management, NSW Agency for Clinical Innovation and the University Seed Grant Fund, School of Health Sciences and Faculty of Medicine and Health. We acknowledge the important work of Dr Bridie Scott-Parker, a pioneer of the BYNDS, a passionate researcher who worked tirelessly in injury prevention research for young drivers.

